# Trends and Intensity of Human Rhinovirus Invasions in Kilifi, Coastal Kenya Over a Twelve-Year Period, 2007-2018

**DOI:** 10.1101/2021.08.03.21261536

**Authors:** John Mwita Morobe, Everlyn Kamau, Nickson Murunga, Winfred Gatua, Martha M. Luka, Clement Lewa, Robinson Cheruiyot, Martin Mutunga, Calleb Odundo, D James Nokes, Charles N. Agoti

## Abstract

**Background:** Human rhinovirus (HRV) is an ubiquitous pathogen and the principal etiologic agent of common cold. Despite the high frequency of HRV infections, data describing its long-term epidemiological patterns in a single population remain limited.

**Methods:** We analysed 1,070 VP4/VP2 genomic region sequences obtained from samples collected between 2007-2018 from hospitalised paediatric patients (< 60 months) with acute respiratory disease in Kilifi County Hospital on the Kenya Coast.

**Results:** Of 7,231 children enrolled, HRV was detected in 1,497 (20.7%) andVP4/VP2 sequences were recovered from 1,070 samples (71.5%). A total of 144 different HRV types were identified (67 HRV-A, 18 HRV-B and 59 HRV-C) and at any time-point, several types co-circulated with alternating predominance. Within types multiple genetically divergent variants were observed. Ongoing HRV infections appeared to be a combination of (i) persistent types (observed up to seven consecutive months), (ii) reintroduced genetically distinct variants and (iii) new invasions (average of 8 new types, annually).

**Conclusion:** Sustained HRV presence in the Kilifi community is mainly due to frequent invasion by new types and variants rather than prolonged circulation of locally established strains.

## Introduction

Human rhinovirus (HRV) is a highly prevalent pathogen and the principal cause of common cold syndrome globally [1,2]. HRV infections result in a wide range of clinical outcomes spanning from asymptomatic and mild illness in the upper airways to severe illness in the lower airways [3,4]. The infections occur in persons of all ages, with the severe presentation more likely in children under the age of 5 years [5,6], the elderly [7] and immunocompromised individuals [8]. Despite the clinical and economic significance of HRV infections, there is little information on the long-term trends and diversity of circulating HRV types.

HRV belongs to the genus *Enterovirus* of the *family Picornaviridae*. The viral single-stranded positive sense RNA genome consists of ∼7200 nucleotides and encodes four structural proteins (VP4, VP2, VP3 and VP1) and seven non-structural proteins (2A, 2B, 2C, 3A, 3B, 3C and 3D) [2]. The three surface-exposed capsid proteins (VP1, VP2 and VP3) carry the antigenically critical sites [9–11]. High genetic variability in the VP4/VP2 and VP1 genomic regions has been instrumental to molecular typing [12,13] and molecular epidemiological investigations[14–16]. Currently, 169 HRV types have been described, and classified into three species i.e., HRV-A, HRV-B and HRV-C (https://www.picornaviridae.com/sg3/enterovirus/enterovirus.htm).

HRV infections occur all year-round in most geographical locations, although peaking in the early autumn and late spring in many temperate countries, and in the rainy season in tropical countries [2,17]. Unclear seasonality and year-round infections have been attributed to lack of inter-type cross-protective immunity [18,19], coupled with the high genetic diversity within the three species, each with the ability to spread independently in a population [14,16,20].

A recent study from Kilifi county located in coastal Kenya spanning 12 months [14] (December 2015 - November 2016), showed that multiple HRV types co-circulate over varied time periods ranging from 1 to 9 months and in most cases each displaying a typical epidemic curve at the local population level; transmission presumably constrained by the decline in susceptibles to that type within the locality. Type-specific (homologous) immunity has been reported to wane after a year [21], and individuals who were previously immune to a particular type gradually become susceptible again to the type again [21,22]. Previous studies found that introduction of new HRV types or sequential invasion by different genetic variants could be due to declining levels of population immunity as well as viral evolution [23,24].

These assertions of perpetually changing HRV types during year-round HRV infections have not been fully investigated in a longitudinal manner [16]. In this study, we analyzed VP4/VP2 sequences of samples collected from hospitalized children with acute respiratory infections (ARIs) between 2007 and 2018 on the Kenyan Coast to evaluate the long-term incidence of the different HRV types, their temporal patterns and intensity of new invasions in a local population.

## Methods

### Study area and population

This study was carried out in Kilifi county, coastal Kenya, which has a population of 1,453,787 and covers an area of ∼12,254km. Nasopharyngeal swab (NPS) samples were obtained from children (<60 months of age) with symptoms of syndromic severe or very severe pneumonia during a paediatric inpatient RSV surveillance study at the county’s referral hospital, Kilifi County Hospital (KCH), between January 2007 and December 2018 [25]. Details of study design, participant recruitment, and sampling procedures have been described elsewhere [25,26]. Ethical approval for the study protocol was obtained from the Scientific and Ethics Review Unit (SERU # 3443) ethics committee, Kenya Medical Research Institute, Nairobi, Kenya.

### HRV screening and sequencing

Viral RNA was extracted and screened for respiratory viruses using a multiplex real-time reverse-transcription PCR (rt-RT-PCR) as described elsewhere [27,28]. A sample was considered HRV positive if rt-RT-PCR cycle threshold (Ct) was <35.0 [26]. A section of VP4/VP2 viral genomic region (∼420 nucleotides long) of positive samples was amplified and sequenced as previously described [14]. Consensus sequences were assembled using the Sequencher software version (Gene Codes Corporation, Ann Arbor, USA).

### Sequence data, HRV species and type assignment

VP4/VP2 sequencing and typing was attempted for all the HRV positive samples collected in 2014, 2016 to 2018. For the years 2010 to 2013 and 2015, 100 HRV positive samples were randomly selected for sequencing proportional to the monthly distribution of positive samples (Supplementary Table 1). Previously published VP4/VP2 sequences from Kilifi (January 2007– December 2009) were retrieved from GenBank (n=271, sequence accession numbers: KY006195 - KY006465) and combined with the 799 newly generated VP4/VP2 sequences (January 2010 – December 2018, GenBank sequence accession numbers; MW622248 - MW623046).

### Definition of terms

We used the term ‘type’ to refer to HRV sequences classified by either cross-neutralization or genetic comparisons as distinct as described previously [13]. Based on this approach, sequences were assigned into the same HRV type based on >90% nucleotide similarity to rhinovirus prototype sequences (also referred to as reference sequences, http://www.picornaviridae.com/sequences/sequences.htm) and phylogenetic clustering with bootstrap support value >70% [13]. Distributions of pairwise genetic distances were assessed for evaluation of intertype and intra-type divergence [13]. Intra-type ‘variant’ was defined on the basis of a divergence threshold value determined as the least frequent value between the first and second modes in a pairwise nucleotide difference distribution plot. Here we are implicitly assuming that sequences with pairwise nucleotide difference falling into the distribution with the low (first) mode are members of the same phylogenetic clade, whereas those with pairwise nucleotide difference within the second distribution with higher mode are members of different phylogenetic clades. A group of viruses within the first, lower distribution were classified as belonging to the same HRV type variant.

The definitions used to describe the temporal occurrence of HRV types are summarised as follows:

a. Persistent: Continued detection, in consecutive or non-consecutive years, of a group of viruses belonging to the same variant of an HRV type.
b. Recurrent: Detection of a virus or group of viruses not observed in the preceding years (>1 year) that belong to a different variant of a previously observed HRV type.
c. Invasion: Detection of a new HRV type not previously locally documented.

### Phylogenetic analyses

Multiple sequence alignments (MSA) were generated using MAFFT v7.220 [29] and maximum likelihood phylogenetic trees estimated using IQ-TREE v1.6.12 [30]. Branch support was assessed by 1000 bootstrap iterations. Temporal signal in the data was examined using TempEst v1.5.3 [31]. To infer time-scaled phylogenies, Bayesian phylogenetic analyses were undertaken in BEAST v.1.10.4 assuming an uncorrelated lognormal relaxed molecular model [32]. The MCMC convergence was assessed in Tracerv1.5 and maximum clade credibility (MCC) trees summarized using TreeAnnotator v1.10.4 with a 10% burn-in. MCC trees were visualized using FigTree v1.4.4.

## Results

### HRV prevalence in Kilifi, 2007 to 2018

Between January 2007 and December 2018, a total of 7,231 NPS samples were collected from children (<60 months of age) admitted with severe pneumonia in KCH (Supplementary Table 1). HRV was detected in 20.7% (1497/7231), with the proportion positive across years ranging from 15.6% to 38.3 % (Supplementary Table 1). The monthly frequency of detection of HRV in the study population is shown in Figure 1. HRV infections were occurring year-round frequently peaking between the month of May and September each year (Figure 1).

**Figure 1.**
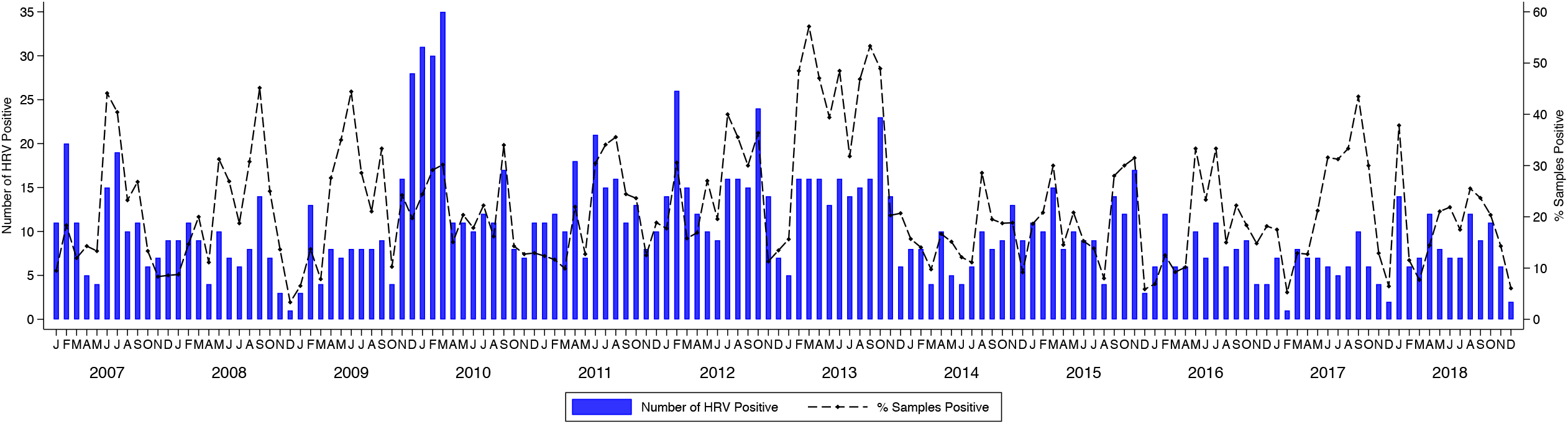
Monthly distribution of human rhinovirus cases identified from surveillance of acute respiratory infections (ARI) in patients under 5 years old admitted to the Kilifi County Hospital, Kenya, 2007–2018. Also included on the secondary y-axis are the proportion (% positivity) of the samples from the in patients with ARI that were HRV positive.

### HRV species and type assignment

A total of 1,070 (71.5%) VP4/VP2 sequences (∼420 nucleotides, some previously reported [26]) were available for this analysis. Of these, 520 (48.6%) sequences were classified as HRV-A comprising 67 distinct HRV-A types; 52 (4.7%) sequences were HRV-B comprising 18 HRV-B types and 498 (46.5%) were HRV-C comprising 59 HRV-C types. HRV-A and HRV-C were more frequently detected while HRV-B infections were low and sporadic (Figure 2(A)). The most commonly detected types were A49 (n = 39), C2 (n = 29), C38 (n = 26), C11 (n = 26), A101 (n=24), A12 (n=23), C6 (n = 22), C21 (n = 21), C3 (n=20) and A78 (n = 19) (Table 1). Twenty four sequences could not be assigned to known HRV types based on the criterion proposed by McIntyre [13] due to these sequences having p-distance of >10.5% with respect to their closest reference sequences (Supplementary Table 2). Other enteroviruses were also detected on sequencing the rt-RT-PCR rhinovirus detections: Enterovirus-D68 (EV-D68) (n=5), human polio virus-1 (n=2), coxsakievirus-B3 (CV-B3) (n=1), coxsakievirus-B2 (CV-B2) (n=1) and echovirus-19 (E-19) (n=1).

**Table 1.** Number of different human rhinovirus types identified in Kilifi, Kenya, 2007 -2018.

| RV-A |  |  |  |  |  |  |  |  |  |  |  |  |  |  |  |
| --- | --- | --- | --- | --- | --- | --- | --- | --- | --- | --- | --- | --- | --- | --- | --- |
| **A49** | **A101** | **A12** | **A78** | **A56** | **A89** | **A20** | **A28** | **A40** | **A54** | **A1** | **A22** | **A61** | **A80** | **A29** | **A30** |
| 39 | 24 | 23 | 19 | 18 | 18 | 17 | 16 | 16 | 14 | 13 | 13 | 13 | 12 | 11 | 10 |
| **A58** | **A63** | **A82** | **A21** | **A75** | **A10** | **A47** | **A65** | **A106** | **A68** | **A103** | **A15** | **A43** | **A81** | **A88** | **A9** |
| 10 | 10 | 10 | 9 | 9 | 8 | 8 | 8 | 7 | 7 | 6 | 6 | 6 | 6 | 6 | 6 |
| **A16** | **A31** | **A46** | **A60** | **A66** | **A73** | **A104** | **A105** | **A13** | **A34** | **A36** | **A45** | **A55** | **A7** | **A90** | **A19** |
| 5 | 5 | 5 | 5 | 5 | 5 | 4 | 4 | 4 | 4 | 4 | 4 | 4 | 4 | 4 | 3 |
| **A24** | **A32** | **A38** | **A39** | **A53** | **A8** | **A94** | **A96** | **A100** | **A102** | **A11** | **A23** | **A67** | **A18** | **A33** | **A41** |
| 3 | 3 | 3 | 3 | 3 | 3 | 3 | 3 | 2 | 2 | 2 | 2 | 2 | 1 | 1 | 1 |
| **A51** | **A59** | **A-**<br>**untyped** |  |  |  |  |  |  |  |  |  |  |  |  |  |
| 1 | 1 | 13 |  |  |  |  |  |  |  |  |  |  |  |  |  |
| RV-B |  |  |  |  |  |  |  |  |  |  |  |  |  |  |  |
| **B4** | **B70** | **B27** | **B42** | **B48** | **B86** | **B91** | **B104** | **B69** | **B102** | **B35** | **B72** | **B83** | **B101** | **B26** | **B6** |
| 7 | 5 | 4 | 4 | 4 | 4 | 4 | 3 | 3 | 2 | 2 | 2 | 2 | 1 | 1 | 1 |
| **B84** | **B92** | **B97** |  |  |  |  |  |  |  |  |  |  |  |  |  |
| 1 | 1 | 1 |  |  |  |  |  |  |  |  |  |  |  |  |  |
| RV-C |  |  |  |  |  |  |  |  |  |  |  |  |  |  |  |
| **C2** | **C38** | **C11** | **C43** | **C6** | **C21** | **C3** | **C10** | **C1** | **C14** | **C22** | **C40** | **C5** | **C27** | **C36** | **C25** |
| 29 | 26 | 26 | 26 | 22 | 21 | 20 | 18 | 17 | 16 | 16 | 16 | 16 | 15 | 13 | 12 |
| **C45** | **C37** | **C31** | **C32** | **C9** | **Cpat19** | **C46** | **Cpat18** | **C12** | **C16** | **C55** | **C15** | **C19** | **C41** | **C51** | **Cpat14** |
| 12 | 11 | 10 | 10 | 10 | 9 | 8 | 8 | 7 | 7 | 6 | 5 | 5 | 5 | 5 | 5 |
| **Cpat21** | **C42** | **Cpat20** | **C23** | **C33** | **C35** | **C39** | **C7** | **C8** | **C26** | **C44** | **C47** | **C49** | **Cpat17** | **Cpat22** | **Cpat28** |
| 5 | 4 | 4 | 3 | 3 | 3 | 3 | 3 | 3 | 2 | 2 | 2 | 2 | 2 | 2 | 2 |
| **C13** | **C17** | **C18** | **C24** | **C29** | **C30** | **C34** | **C48** | **C50** | **Cpat16** | **Cpat27** | **C-**<br>**untyped** |  |  |  |  |
| 1 | 1 | 1 | 1 | 1 | 1 | 1 | 1 | 1 | 1 | 1 | 11 |  |  |  |  |

**Figure 2.**
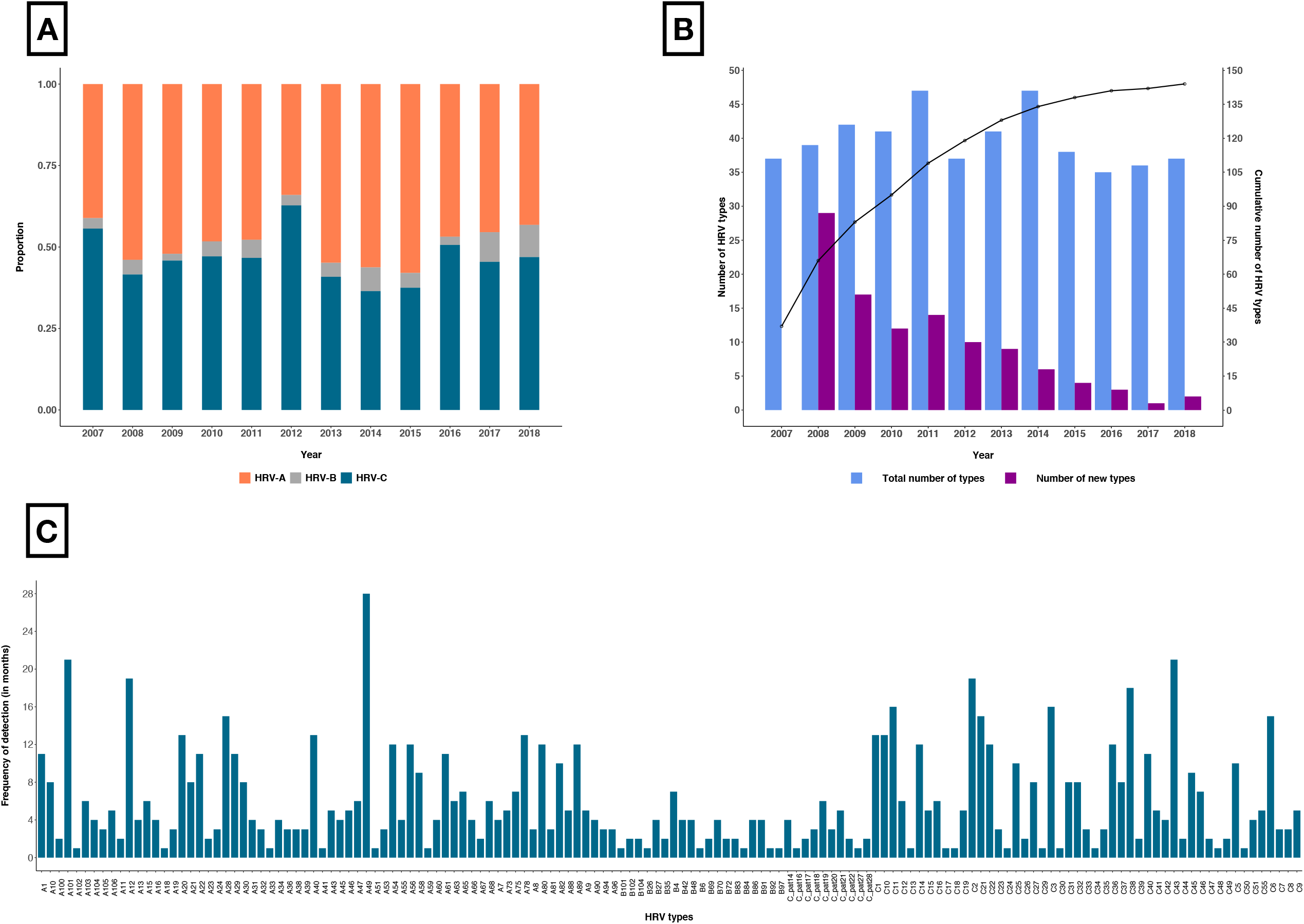
(A) Annual proportion of HRV species across the 12-years period. (B)The total number (blue bars) and new number (purple bars) of HRV types detected annually over the period 2007-2018. Also shown is the cumulative number of all the different HRV types observed during the study period (black line). (C) The overall frequency of detection in months or the number of months each HRV type was detected. The types are ordered alphabetically.

### Temporal trends of HRV types in Kilifi

We detected on average, 39 HRV types annually (range, 35 -47), a mean of 8 (range, 1-29) of which were new HRV types identified for the first time in the population each year from 2008 as others previously detected types disappeared (Figure 2(B)). The cumulative number of new HRV types detected annually increased rapidly since the beginning of the surveillance period and then saturated after approximately 9 years (Figure 2(B)). HRV types commonly co-circulated and with varying frequency in the 12-year period (Figure 2(C), Supplementary File 1). Several types were present at high prevalence while others occurred once or sporadically. Some types circulated consecutively for months: for example, A56 was detected in 7 consecutive months (May to November 2007), C11 was present for 6 consecutive months (February to July 2016), and C38, A40 and C2 types circulated consecutively for 5 months (November 2009 to March 2010, April to August 2016 and May to September 2010, respectively) (Supplementary File 1).

Additionally, several types recurred after considerable periods of absence. For example, A12 first seen in February 2007 was not detected again until February 2009, 23 months later, while C38 viruses were detected 4 years apart between 2012 and 2016 (Figure 3, Supplementary File 1). Temporally, several HRV types exhibited synchronized co-circulation and recurrence, for example (i) C1, C11, C2, C38, C22 and C21; (ii) A75, A89, A12, A28, A96, A106, A80 and A10; (iii) A90, A55, A61, A45, A54 and A60; (iv) C14, C41, C45, C10, C16, C25, C32 and C47.

**Figure 3.**
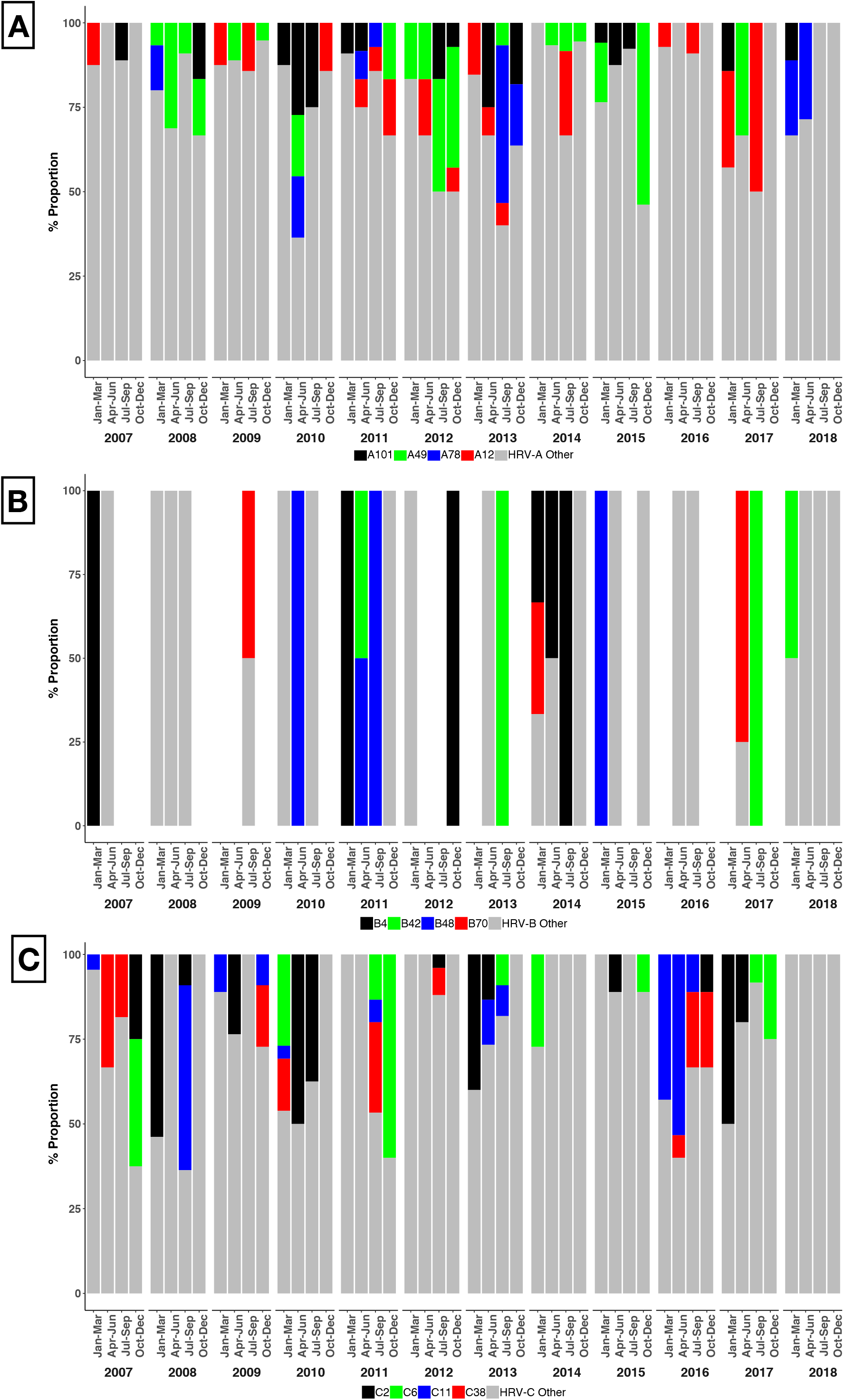
Quarterly proportions of HRV types detected organised at the species level; shown here are the temporal trends of 5 most prevalent types per species while the rest are indicated as others. A) Quarterly proportion of HRV A types (B) Quarterly proportion of HRV B types, (C) Quarterly proportion of HRV C types.

### Genetic diversity of HRV types in Kilifi

The nucleotide (nt) sequence identity among HRV-A, -B and -C viruses was determined as 57.3–100%, 66.0–100%, and 45.1–100%, respectively, and 59.8– 100%, 79.9–100% and 53.3–100% at the amino acid (aa) level, respectively. Intra-type nucleotide variation was observed in the VP4/VP2 region of viruses sampled over the 12 years study period (Figure 4(A), Supplementary Figure 1). Nonetheless, the substitutions were mostly synonymous i.e., not amino acid changing. The distribution of pairwise nucleotide distances showed multi-modal peaks suggesting circulation of distinct variants within individual HRV types (Figure 4(B), Supplementary Figure 2). These observations were congruent with multiple within-type phylogenetic clusters.

**Figure 4.**
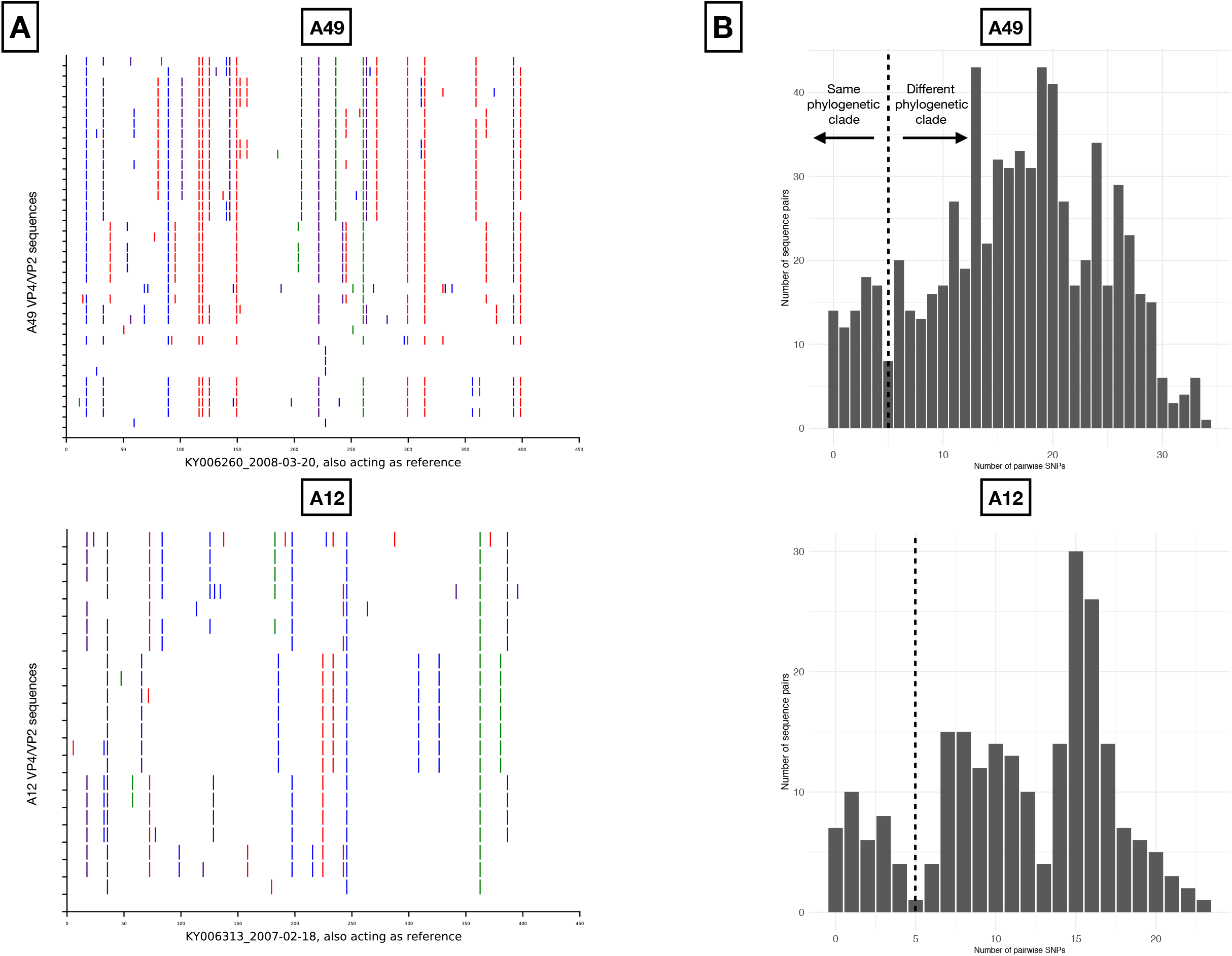
(A) Nucleotide variability across the sequenced VP4/VP2 region for types A49 and C2. For each type, the viruses were compared to the earliest sampled sequence. Vertical coloured bars show the nucleotide differences: red is a change to T, orange is a change to A, purple is a change to C and blue is a change to G. (B) Distribution of pairwise nucleotide difference for the VP4/VP2 region of types A49 and C2.

Several HRV types were characterised by genetically distinct temporal clusters, for example, A49, C38 and A101 (Figure 5). A49 was detected as eleven distinct variants circulating at different periods, three of which occurred as singletons (single sequences) suggesting under sampled genetic diversity (Figure 5, Table 2). Multiple genetic variants of type C6 co-circulated during 2010/ 2011 (Supplementary Figure 3, Table 2), which likely indicates separate virus introductions into the Kilifi population. Several HRV types had variants that contained sequences from multiple years indicating variant persistence over an extended period or repeated reintroductions, e.g., A101 variant 5 (v5) comprised of viruses observed from 2010 to 2013 (Figure 5, Table 2).

**Figure 5.**
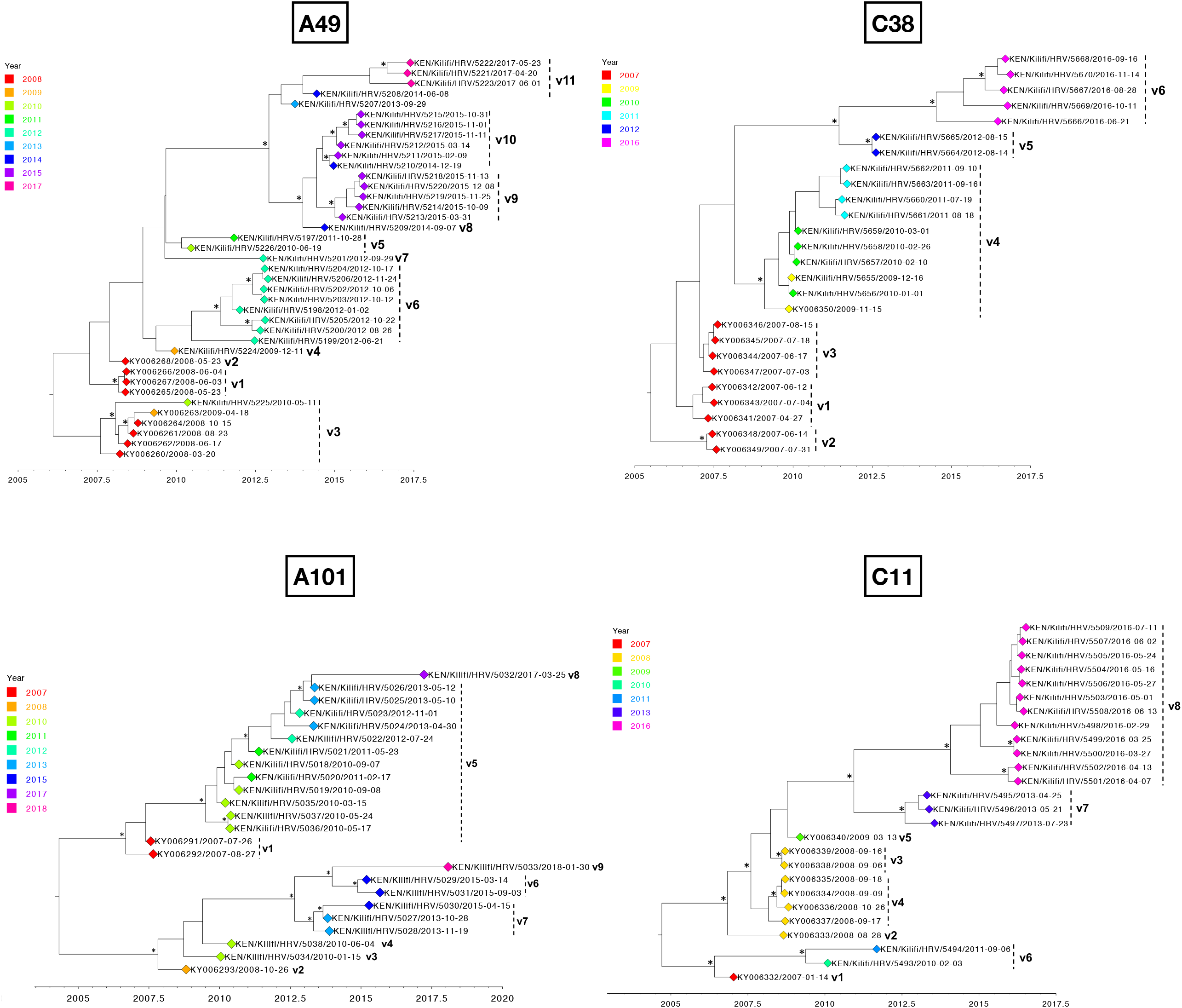
Bayesian phylogenetic trees showing the VP4/VP2 region of the HRV types A49, C2, C38 and C11.Variant names are next to the phylogenetic clusters, e.g., v1 representing variant 1 for a specific type. Node support is indicated by (*) for posterior probabilities > 0.9.

**Table 2.** Number of variants for the ten most prevalent HRV types identified in Kilifi, Kenya, 2007 to 2018; * where b in a, b are the singletons (single sequence which genetically distinct from other types).

| HRV type | Number of<br>sequences | Number of variants,<br>singletons | Period (Year) |
| --- | --- | --- | --- |
| A12 | 23 | 9,5 | v1(2007), v2(2007), v3(2009), v4(2009-2011), v5(2014),<br>v6(2014), v7(2016), v8(2016), v9(2017) |
| A78 | 19 | 7,3 | v1(2008), v2(2010-2011), v3(2011), v4(2013), v5(2013),<br>v6(2018), v7(2018) |
| C2 | 29 | 12,5 | v1(2007-2008), v2(2008), v3(2008), v4(2009), v5(2009),<br>v6(2010), v7(2010), v8(2012), v9(2013), v10(2015), v11(2016-2017), v12(2017) |
| C11 | 25 | 8,3 | v1(2007), v2(2008), v3(2008), v4(2008), v5(2009), v6(2010-2011), v7(2013), v8(2016) |
| C21 | 21 | 6,2 | v1(2007), v2(2007-2012), v3(2010), v4(2012), v5(2015),<br>v6(2018) |
| C38 | 26 | 6,0 | v1(2007), v2(2007), v3(2007), v4(2009-2011), v5(2012) ,<br>v6(2016) |
| C3 | 20 | 7,2 | v1(2009), v2(2010 -201), v3(2011), v4(2013), v5(2015),<br>v6(2017), v7(2018) |
| A49 | 39 | 11,4 | v1(2008), v2(2008), v3(2008-2010), v4(2009), v5(2010-2011),<br>v6(2012), v7(2012), v8(2014), v9(2015), v10(2014-2015),<br>v11(2013-2014,2017) |
| A101 | 25 | 9,5 | v1(2007), v2(2008), v3(2010), v4(2010), v5(2010-2013),<br>v6(2015), v7(2013-2015), v8(2017), v9(2018) |
| C6 | 22 | 11,4 | v1(2007), v2(2010 - 2011), v3(2010), v4(2010), v5(2010),<br>v6(2011), v7(2011), v8(2011), v9(2013-2014), v10(2015),<br>v11(2017) |

## Discussion

We describe HRV molecular epidemiology in Kilifi over a 12-year period (2007 to 2018). Our study documents a long-term pattern of co-circulation, persistence and invasion of types in this rural community of coastal Kenya. Consistent with other previous studies, HRV was ubiquitous at the population scale and multiple types co-circulated even within a single month [14,16,20]. Among the HRV cases detected, HRV-B was least frequently detected. It is not clear why HRV-B is less diverse and each type within it was on average less frequent. The observed proportions of HRV types are consistent with similar studies elsewhere [16,33].

The majority (99%) of our sequences occurred within the proposed divergence thresholds for HRV typing and classification using VP4/VP2 region (10.5% for HRV-A, 9.5% for HRVB, and 10.5% for HRV-C) [13]. This exemplifies significant sequence conservation in the VP4/VP2 region within a type allowing robust genotypic assignment. However, 24 sequences did not fit the classification system for VP4/VP2 region and require further investigation such as by whole genome sequencing to also consider variation in the VP1 region to determine whether these sequence correspond to new types [13].

In total, 144 distinct HRV types were detected over the 12-year period, representing 85% of the currently known types (http://www.picornaviridae.com). The finding of enteroviruses (EV) sequences reflects the cross-reactivity of the HRV detection PCR for non-rhinovirus EV, due to their close relationship and the nucleotide sequence conservation of the 5’-untranslated region that is targeted by our diagnostic assay [34,35]. Half of the EVs (5/10) were EV-D68, an EV that is also associated with respiratory disease [36]. Two sequences were found to be HPV-1. The children of 23-and 10-months age had probably been immunized (oral polio vaccine) just before the nasopharyngeal swab. The other EVs detected (CV-B3, CV-B2 and E-19) are found only occasionally in respiratory samples [37].

Unique types (never seen previously from the start of surveillance in 2007) invaded the population every year during the surveillance period. The frequent invasions could be explained by lack of pre-existing immune memory or weak heterotypic immunity [38]. Recurrence of HRV types could be promoted by antigenic variation occurring on the other surface proteins (VP2, VP1 and VP3) allowing infection where prior infection confers only incomplete or short-lived immunity to future evolved variants. Recurrence, particularly where the recurring strains were genetically identical to older circulating strains, may also be observed in the portion of the population that had not been previously exposed to a HRV type.

Some HRV types were seen once or very sporadically and could be associated with mild disease or asymptomatic infections or have reduced transmission rates probably suppressed by pre-existing host population immunity [39]. The number of new types detected in the paediatric hospitalizations decreased over time, with the cumulative number of types identified levelling off in 2016, perhaps indicating the period a given study cohort will take to experience every known HRV type or maximum number of types.

In some HRV types, VP4/VP2 coding region remained conserved at both nucleotide and amino acid level after periods of quiescence, which probably ensures strain survival by maintaining low-level genetic variation. In a linear strain space, strains interact via cross-immunity to nearby strains with shared epitopes and this interaction tails off with genetic distance [40]. Yet, the VP4/VP2 region might not be primarily antigenic [41], and genetic changes could have occurred at immunogenic sites located in other capsid proteins (VP1 or VP3). Genome-wide sequence data would therefore be useful to confirm strain conservation and maintenance. The evident intra-type genetic diversity with differential temporal distribution could suggest sequential virus introductions or diversification of locally circulating variants [42].

This study had some limitations. First, in some years (2010 to 2013 and 2015) we only sequenced a proportion (∼72%) of the positive cases, this might underestimate the diversity and rhinovirus types circulating in the population. Samples selected were prioritised on the basis of sample viral load as indicated by Ct value and monthly distribution of samples. Second, we only sequenced the VP4/VP2 coding region, but the more reliable phylogenetic relationships will be defined from full-length genome analysis [43]. Future studies with whole-genome data are required to give improved understanding on the molecular epidemiology of HRV.

In conclusion, this study describes the nature of HRV infections within a community: repeated invasion by heterogeneous HRV types, rather than long-term continuity of the same HRV types, as well as continuous diversification of circulating variants, and enhances our understanding on rhinovirus transmission dynamics at the population level.

## Data Availability

All data generated and analysis script for this manuscript are available from the Virus Epidemiology and Control (VEC), KEMRI-Wellcome Trust Research Programme, data server under the DOI: https://doi.org/10.7910/DVN/OL399P

## Acknowledgements

We thank the study participants for providing the study samples. We also thank all members of the Virus Epidemiology and Control (VEC) Research Group in Kilifi who were involved at various stages of this study. We also thank the parents and guardians of the children for accepting to participate in this study.

## Author Contributions

J.M.M, E.K., C.A., D.J.N., designed the study, J.M.M., C.L., R.C., M.M., C.O. performed HRV screening and sequencing, J.M.M., E.K., M.M.L, N.M., curated and analysed data and J.M.M., E.K., C.A., D.J.N., wrote the manuscript. All authors have read and approved the final manuscript draft.

## Supplementary Materials

**Supplementary File 1** Monthly distribution and frequency of all the 144 types detected in Kilifi, Kenya, between 2007 and 2018.

**Supplementary Table 1** Summary of the number of samples tested, samples positive for HRV, and sequences obtained from pediatric patients (< 60 months old) admitted at the Kilifi County Hospital with acute respiratory illness.

**Supplementary Table 2** Untyped VP4/VP2 sequences and p-distance to the closest HRV reference sequence.

**Supplementary Figure 1** Nucleotide variability across the sequenced VP4/VP2 region for types A101, C6, C2, C38, A78, C11, C21 and C3. For each type, the viruses were compared to the earliest sampled sequence. Vertical coloured bars show the nucleotide differences: red is a change to T, orange is a change to A, purple is a change to C and blue is a change to G.

**Supplementary Figure 2** Distribution of pairwise nucleotide difference for the VP4/VP2 region of types A101, C6, C2, C38, A78, C11, C21 and C3.

**Supplementary Figure 3** Bayesian phylogenetic trees showing the VP4/VP2 region of the HRV types A101, A12, C6, C21, C3 and A78. Variant names are next to the phylogenetic clusters, e.g., v1 representing variant 1 for the various types. Node support is indicated b (*) for posterior probabilities > 0.9

## References

1. Ouédraogo S, Traoré B, Nene Bi ZAB, et al. Viral Etiology of Respiratory Tract Infections in Children at the Pediatric Hospital in Ouagadougou (Burkina Faso). PLoS One. 2014 Oct;9(10):e110435.

2. Jacobs SE, Lamson DM, Kirsten S, Walsh TJ. Human rhinoviruses. Clin Microbiol Rev. 2013 Jan;26(1):135–62.

3. Fry AM, Lu X, Olsen SJ, et al. Human Rhinovirus Infections in Rural Thailand: Epidemiological Evidence for Rhinovirus as Both Pathogen and Bystander. PLoS One. 2011 Mar;6(3):e17780.

4. van Benten I, Koopman L, Niesters B, et al. Predominance of rhinovirus in the nose of symptomatic and asymptomatic infants. Pediatr Allergy Immunol. 2003 Oct;14(5):363–70.

5. Miller EK, Lu X, Erdman DD, et al. Rhinovirus-associated hospitalizations in young children. J Infect Dis. 2007;195.

6. Luka MM, Kamau E, Adema I, et al. Molecular epidemiology of human rhinovirus from one-year surveillance within a school setting in rural coastal Kenya. medRxiv. 2020 Jan 1;2020.03.09.20033019.

7. Hung IFN, Zhang AJ, To KKW, et al. Unexpectedly Higher Morbidity and Mortality of Hospitalized Elderly Patients Associated with Rhinovirus Compared with Influenza Virus Respiratory Tract Infection. Int J Mol Sci. 2017 Feb;18(2).

8. Piralla A, Zecca M, Comoli P, Girello A, Maccario R, Baldanti F. Persistent rhinovirus infection in pediatric hematopoietic stem cell transplant recipients with impaired cellular immunity. J Clin Virol. 2015;67:38–42.

9. Sherry B, Rueckert R. Evidence for at least two dominant neutralization antigens on human rhinovirus 14. J Virol. 1985 Jan;53(1):137–43.

10. Sherry B, Mosser AG, Colonno RJ, Rueckert RR. Use of monoclonal antibodies to identify four neutralization immunogens on a common cold picornavirus, human rhinovirus 14. J Virol. 1986 Jan;57(1):246—257.

11. Stepanova E, Isakova-Sivak I, Rudenko L. Overview of human rhinovirus immunogenic epitopes for rational vaccine design. Expert Rev Vaccines. 2019 Sep;18(9):877–80.

12. Simmonds P, McIntyre C, Savolainen-Kopra C, Tapparel C, Mackay IM, Hovi T. Proposals for the classification of human rhinovirus species C in genotypically assigned types. J Gen Virol. 2010;91.

13. McIntyre CL, Knowles NJ, Simmonds P. Proposals for the classification of human rhinovirus species A, B and C into genotypically assigned types. J Gen Virol. 2013 Aug;94(Pt 8):1791–806.

14. Morobe JM, Nyiro JU, Brand S, et al. Human rhinovirus spatial-temporal epidemiology in rural coastal Kenya, 2015-2016, observed through outpatient surveillance. Wellcome Open Res. 2019;3(128).

15. Arakawa M, Okamoto-Nakagawa R, Toda S, et al. Molecular epidemiological study of human rhinovirus species A, B and C from patients with acute respiratory illnesses in Japan. J Med Microbiol. 2011/10/22. 2012;61(Pt 3):410–9.

16. van der Linden L, Bruning AH, Thomas X V, et al. A molecular epidemiological perspective of rhinovirus types circulating in Amsterdam from 2007 to 2012. Clin Microbiol Infect. 2016/08/25. 2016;22(12):1002.e9-1002.e14.

17. Garcia J, Espejo V, Nelson M, et al. Human rhinoviruses and enteroviruses in influenza-like illness in Latin America. Virol J. 2013 Oct;10:305.

18. Glanville N, Johnston SL. Challenges in developing a cross-serotype rhinovirus vaccine. Curr Opin Virol. 2015 Apr;11:83–8.

19. Glanville N, McLean GR, Guy B, et al. Cross-serotype immunity induced by immunization with a conserved rhinovirus capsid protein. PLoS Pathog. 2013;9(9):e1003669.

20. Sansone M, Andersson M, Brittain-Long R, Andersson LM, Olofsson S, Westin J, Lindh M. Rhinovirus infections in western Sweden: a four-year molecular epidemiology study comparing local and globally appearing types. Eur J Clin Microbiol Infect Dis. 2013/02/26. 2013;32(7):947–54.

21. Barclay WS, al-Nakib W, Higgins PG, Tyrrell DA. The time course of the humoral immune response to rhinovirus infection. Epidemiol Infect. 1989 Dec;103(3):659–69.

22. Barclay WS, Callow KA, Sergeant M, al-Nakib W. Evaluation of an enzyme-linked immunosorbent assay that measures rhinovirus-specific antibodies in human sera and nasal secretions. J Med Virol. 1988 Aug;25(4):475–82.

23. Jartti T, Lee W-M, Pappas T, Evans M, Lemanske Jr RF, Gern JE. Serial viral infections in infants with recurrent respiratory illnesses. Eur Respir J. 2008/04/30. 2008 Aug;32(2):314–20.

24. van der Zalm MM, Wilbrink B, van Ewijk BE, Overduin P, Wolfs TFW, van der Ent CK. Highly frequent infections with human rhinovirus in healthy young children: A longitudinal cohort study. J Clin Virol. 2011;52:317–20.

25. Nokes DJ, Ngama M, Bett A, et al. Incidence and severity of respiratory syncytial virus pneumonia in rural Kenyan children identified through hospital surveillance. Clin Infect Dis. 2009 Nov 1;49(9):1341–9.

26. Onyango CO, Welch SR, Munywoki PK, et al. Molecular epidemiology of human rhinovirus infections in Kilifi, coastal Kenya. J Med Virol. 2012 May;84(5):823–31.

27. Gunson RN, Collins TC, Carman WF. Real-time RT-PCR detection of 12 respiratory viral infections in four triplex reactions. J Clin Virol. 2005 Aug;33(4):341–4.

28. Hammitt LL, Kazungu S, Welch S, et al. Added Value of an Oropharyngeal Swab in Detection of Viruses in Children Hospitalized with Lower Respiratory Tract Infection. J Clin Microbiol. 2011 Jun 1;49(6):2318 LP – 2320.

29. Katoh K, Standley DM. MAFFT multiple sequence alignment software version 7: improvements in performance and usability. Mol Biol Evol. 2013 Apr;30(4):772–80.

30. Nguyen L-T, Schmidt HA, von Haeseler A, Minh BQ. IQ-TREE: A Fast and Effective Stochastic Algorithm for Estimating Maximum-Likelihood Phylogenies. Mol Biol Evol. 2015 Jan 1;32(1):268–74.

31. Rambaut A, Lam TT, Max Carvalho L, Pybus OG. Exploring the temporal structure of heterochronous sequences using TempEst (formerly Path-O-Gen). Virus Evol. 2016 Jan;2(1).

32. Li WLS, Drummond AJ. Model averaging and Bayes factor calculation of relaxed molecular clocks in Bayesian phylogenetics. Mol Biol Evol. 2011/09/22. 2012 Feb;29(2):751–61.

33. Esposito S, Daleno C, Baggi E, et al. Circulation of different rhinovirus groups among children with lower respiratory tract infection in Kiremba, Burundi. Eur J Clin Microbiol Infect Dis Off Publ Eur Soc Clin Microbiol. 2012 Nov;31(11):3251–6.

34. Tapparel C, Cordey S, Van Belle S, et al. New molecular detection tools adapted to emerging rhinoviruses and enteroviruses. J Clin Microbiol. 2009 Jun;47(6):1742–9.

35. Oberste MS, Nix WA, Maher K, Pallansch MA. Improved molecular identification of enteroviruses by RT-PCR and amplicon sequencing. J Clin Virol. 2003;26(3):375–7.

36. Xiang Z, Wang J. Enterovirus D68 and Human Respiratory Infections. Semin Respir Crit Care Med. 2016/08/03. 2016 Aug;37(4):578–85.

37. Royston L, Tapparel C. Rhinoviruses and Respiratory Enteroviruses: Not as Simple as ABC. Viruses. 2016/01/14. 2016;8(1).

38. Rathe JA, Liu X, Tallon LJ, Gern JE, Liggett SB. Full-genome sequence and analysis of a novel human rhinovirus strain within a divergent HRV-A clade. Arch Virol. 2010;155(1):83–7.

39. Hamilton W, Weiss RA, Wain–Hobson S, May RM, Gupta S, McLean AR. Infectious disease dynamics: what characterizes a successful invader? Philos Trans R Soc London Ser B Biol Sci. 2001 Jun;356(1410):901–10.

40. Gog JR, Grenfell BT. Dynamics and selection of many-strain pathogens. Proc Natl Acad Sci. 2002 Dec;99(26):17209 LP – 17214.

41. Ledford RM, Patel NR, Demenczuk TM, Watanyar A, Herbertz T, Collett MS, Pevear DC. VP1 sequencing of all human rhinovirus serotypes: insights into genus phylogeny and susceptibility to antiviral capsid-binding compounds. J Virol. 2004/03/16. 2004;78(7):3663–74.

42. Agoti CN, Otieno JR, Ngama M, Mwihuri AG, Medley GF, Cane PA, Nokes DJ. Successive Respiratory Syncytial Virus Epidemics in Local Populations Arise from Multiple Variant Introductions, Providing Insights into Virus Persistence. J Virol. 2015 Nov;89(22):11630–42.

43. Luka MM, Kamau E, de Laurent ZR, Morobe JM, Alii LK, Nokes DJ, Agoti CN. Whole genome sequencing of two human rhinovirus A types (A101 and A15) detected in Kenya, 2016-2018. Wellcome Open Res 2021 6178. 2021 Jul 8 ;6:178.

