## Supplementary material for "Trends and Intensity of Human Rhinovirus Invasions in Kilifi, Coastal Kenya Over a Twelve-Year Period, 2007-2018": Supplimentary Table 1

**Supplementary Table 1:** Summary of the number of samples tested, samples positive for HRV, and sequences obtained from pediatric patients (≤ 60 months old) admitted at the Kilifi County Hospital with acute respiratory illness.

| Year | Hospital admissions | Samples  tested | Samples  Positives for HRV  (%) | Number of sequences (%) |
| --- | --- | --- | --- | --- |
| 2007 | 1174 | 569 | 128 (22.5) | 124 (96.9) |
| 2008 | 1001 | 429 | 89 (20.8) | 87 (97.8) |
| 2009 | 1103 | 526 | 116 (22.1) | 96 (82.8) |
| 2010 | 1084 | 897 | 190 (21.2) | 87 (44.8) |
| 2011 | 905 | 805 | 148 (18.4) | 90 (60.8) |
| 2012 | 874 | 787 | 172 (21.9) | 94 (54.7) |
| 2013 | 583 | 444 | 170 (38.3) | 93 (54.7) |
| 2014 | 815 | 619 | 106 (17.1) | 96 (90.6) |
| 2015 | 806 | 629 | 120 (19.1) | 88 (73.3) |
| 2016 | 791 | 572 | 89 (15.6) | 79 (88.7) |
| 2017 | 535 | 367 | 68 (18.5) | 55 (80.9) |
| 2018 | 856 | 587 | 101(17.2) | 81 (80.2) |
| Total | 10527 | 7231 | 1497 (20.7) | 1070 (71.5) |
