## Supplementary material for "Trends and Intensity of Human Rhinovirus Invasions in Kilifi, Coastal Kenya Over a Twelve-Year Period, 2007-2018": Supplimentary Table 2

**Supplementary Table 2** Untyped VP4/VP2 sequences and *p*-distance to the closest HRV reference sequence.

| Sequence ID | Closest HRV Type (p-distance) |
| --- | --- |
| KY006312 | HRV-A20 (16.0) |
| KY006259 | HRV-A30 (14.2) |
| KY006269 | HRV-A30 (16.1) |
| KY006220 | HRV-A40 (12.1) |
| KEN/Kilifi/HRV/5169/2011-06-27 | HRV-A40 (12.4) |
| KY006280 | HRV-A58 (12.1) |
| KEN/Kilifi/HRV/5548/2015-08-09 | HRV-A7 (11.4) |
| KEN/Kilifi/HRV/5554/2015-09-06 | HRV-A7 (11.4) |
| KY006279 | HRV-A7 (12.4) |
| KY006278 | HRV-A7 (13.6 |
| KY006321 | HRV-A8 (13.3) |
| KY006320 | HRV-A8 (14.4) |
| KY006290 | HRV-A80 (13.8) |
| KY006452 | HRV-C10 (12.4) |
| KEN/Kilifi/HRV/5747/2018-05-09 | HRV-C27 (10.6) |
| KEN/Kilifi/HRV/5748/2018-05-10 | HRV-C27 (10.6) |
| KEN/Kilifi/HRV/5387/2013-10-24 | HRV-C27 (14.4) |
| KEN/Kilifi/HRV/5457/2014-08-30 | HRV-C28 (13.6) |
| KEN/Kilifi/HRV/5721/2018-01-12 | HRV-C8 (12.4) |
| KEN/Kilifi/HRV/5725/2018-01-26 | HRV-C8 (12.4) |
| KEN/Kilifi/HRV/5728/2018-02-06 | HRV-C8 (12.4) |
| KEN/Kilifi/HRV/5729/2018-02-15 | HRV-C8 (14.4) |
| KEN/Kilifi/HRV/5740/2018-04-16 | HRV-C9 (22.2) |
| KEN/Kilifi/HRV/5741/2018-04-20 | HRV-C9 (22.2) |
