## Supplementary figures and images for "Trends and Intensity of Human Rhinovirus Invasions in Kilifi, Coastal Kenya Over a Twelve-Year Period, 2007-2018"

### Supplimentary Figure 1

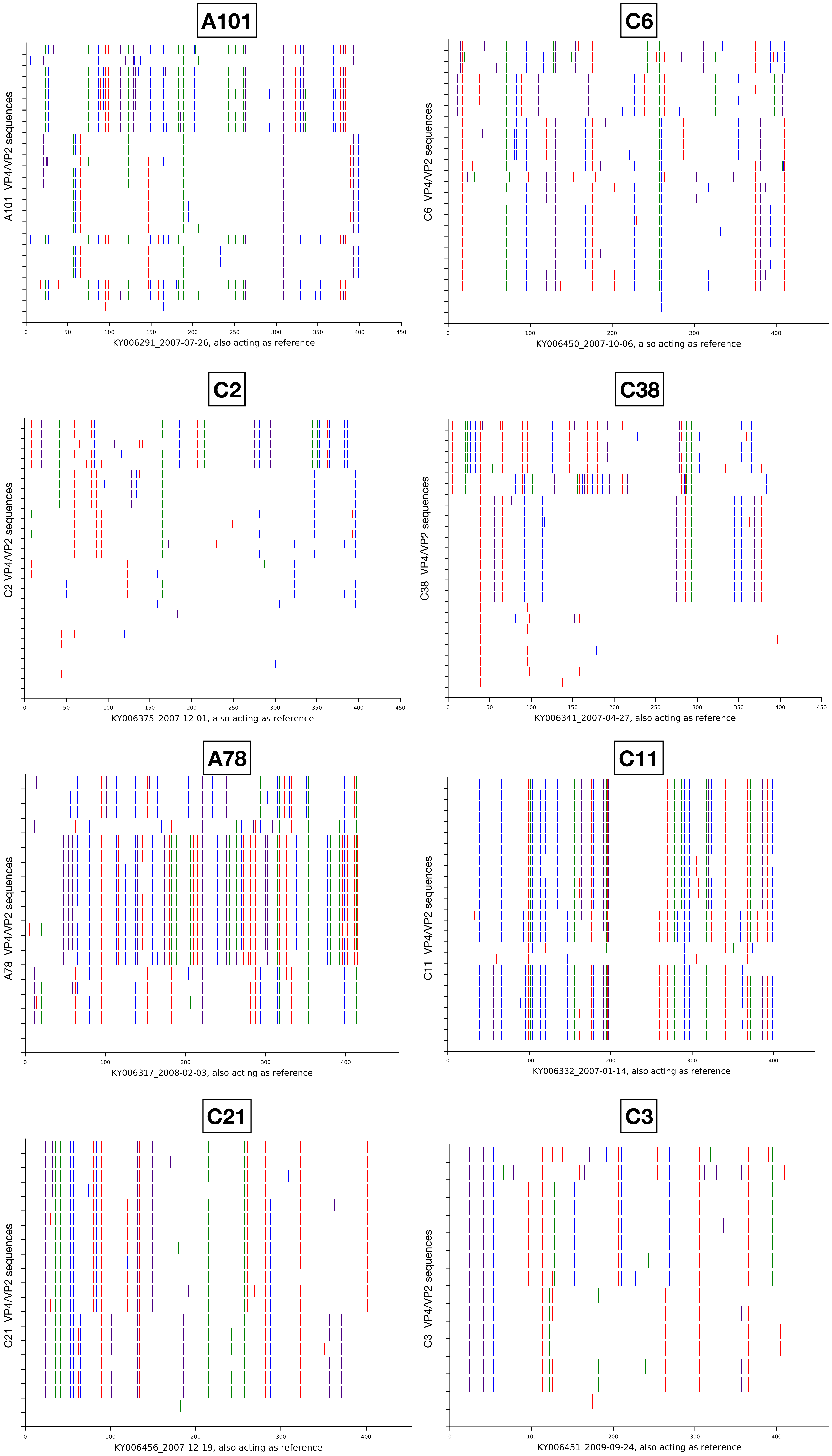

### Supplimentary Figure 2

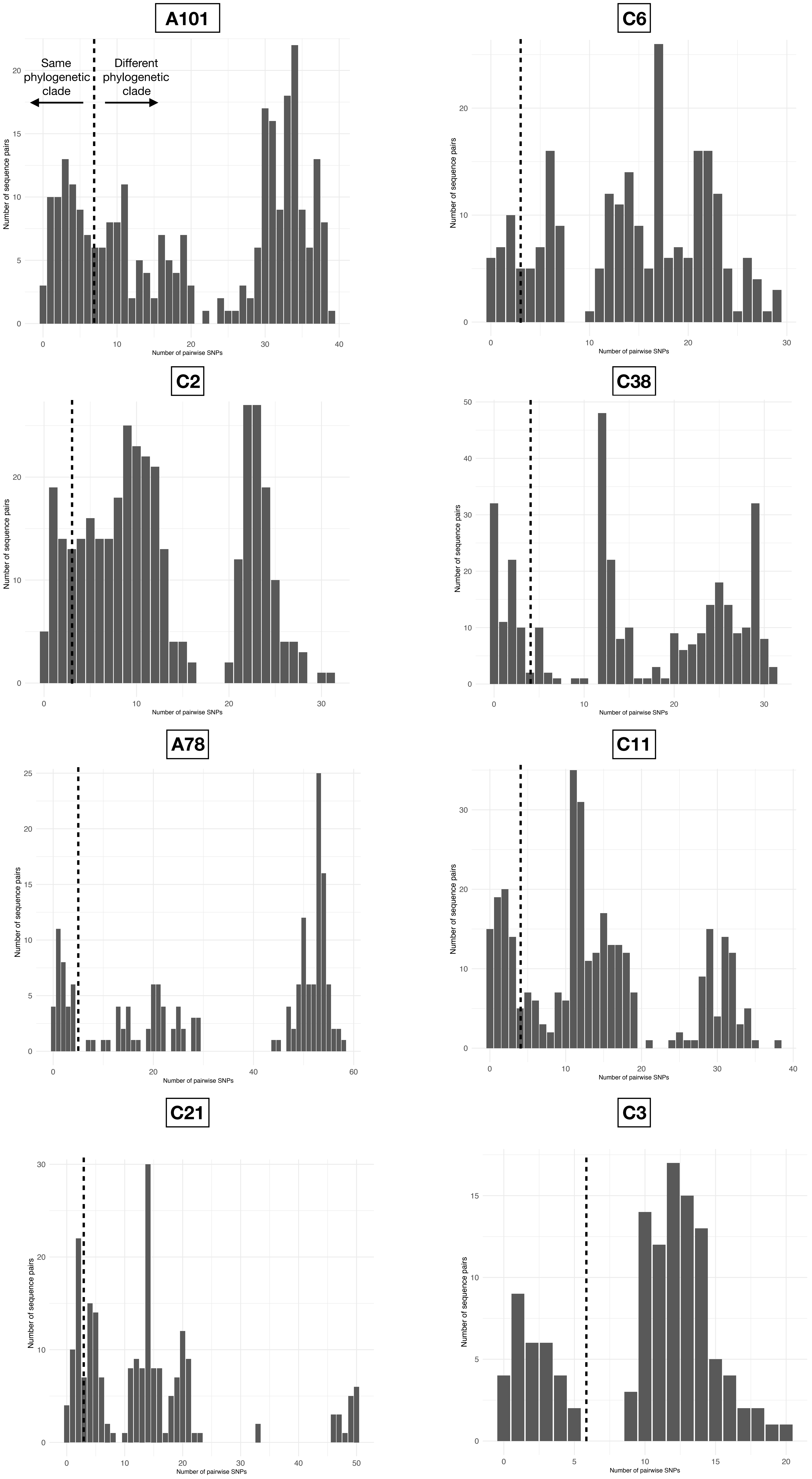

### Supplimentary Figure 3

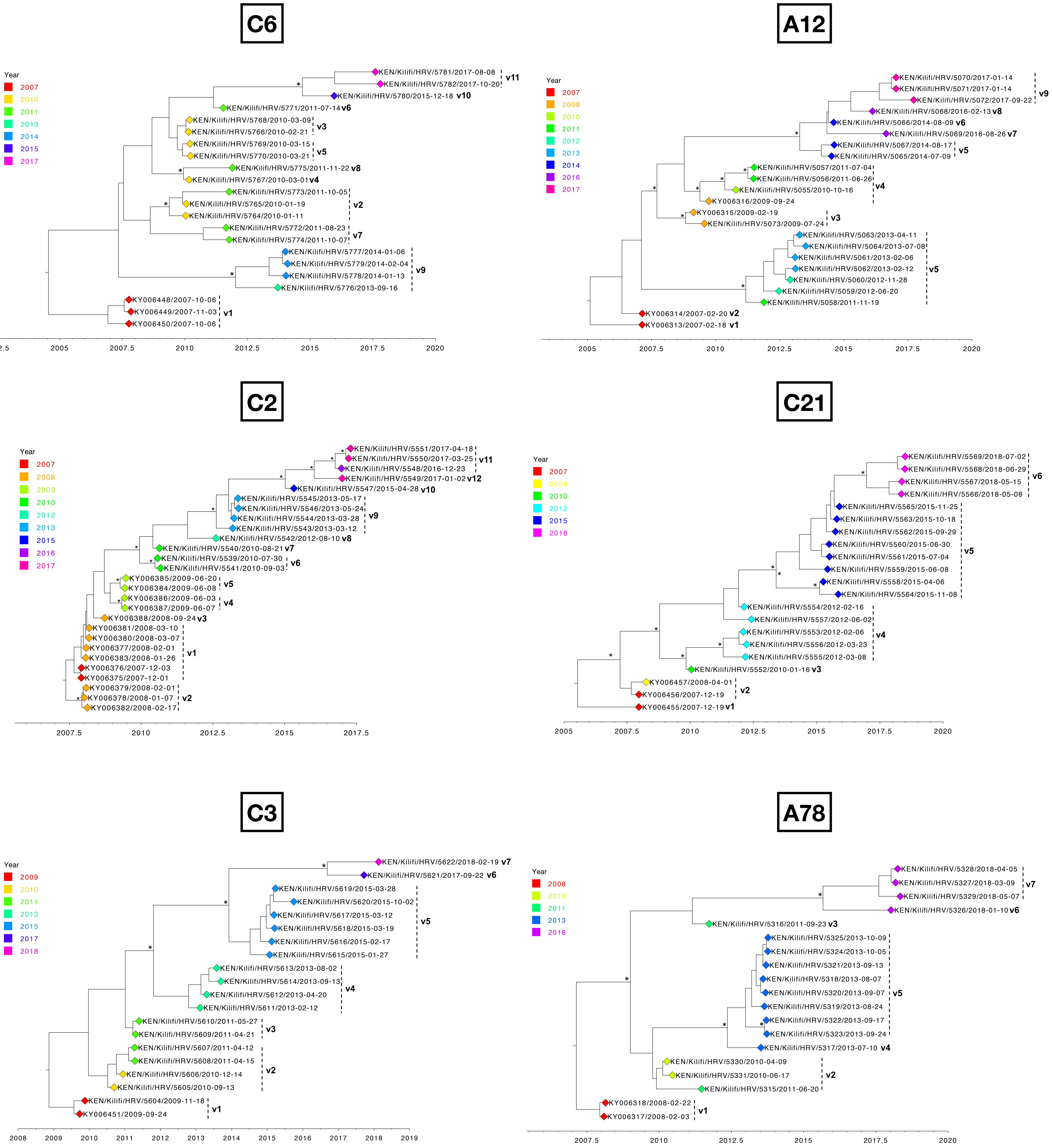
